# Severe bandemia is not associated with increased risk for adverse events in general pediatric emergency department patients

**DOI:** 10.1101/2020.12.28.20248556

**Authors:** Daniel Najafali, Emilie Berman, Tiffany T Cao, Allison Karwoski, Norvir Kaur, Ikram Afridi, Norhan Abdalla, Leenah Afridi, Iana Sahadzic, Juliana Solomon, Isha Yardi, Quincy K Tran

**Affiliations:** University of Maryland School of Medicine, The Research Associate Program in Emergency Medicine and Critical Care, Baltimore, MD, USA; University of Maryland School of Medicine, Department of Emergency Medicine, Baltimore, MD, USA; University of Maryland School of Medicine, The R Adams Cowley Shock Trauma Center, Baltimore, MD, USA

**Keywords:** Adverse outcome, Bacterial illness, Bandemia, General pediatric emergency department patients, Severe bandemia

## Abstract

**Background:** The presence of band cells >10% of the total white blood cell (WBC) count (“bandemia”) is often used as an indicator of serious bacterial illness (SBI). Results from studies of bandemia as a predictor of SBI were conflicting and little is known about the relationship between severe bandemia (SB) and clinical outcomes from SBI in children.

**Objectives:** In this retrospective patient-centered study, we hypothesized that SB (band level >20%) is not associated with adverse outcomes in an emergency department (ED) pediatric population.

**Methods:** Medical records from children between the ages of 2 months and 18 years with SB who presented to a tertiary referral regional hospital were studied. Outcomes were categorized as severe adverse events (SAEs) or moderate adverse events (MAEs). Multivariate logistic regressions were used to assess the association between SB and outcomes.

**Results:** We analyzed 102 patients. Mean age (standard deviation [SD]) was 5.25(0.5) years, 18 (18%) had MAE, 21 (21%) had SAE, and no patients died. Mean band levels were similar between groups: no adverse events [28 (10)] vs. SAE [31 (9)] vs. MAE [27(8)], p=0.64. Multivariate logistic regressions showed SB was not associated with any adverse events (odds ratio [OR] 1.04, 95% confidence interval [CI] 0.9-1.1, p=0.27). Non-normal X-ray (XR) (OR 17, 95% CI 3.3-90, p<0.001) was associated with MAE, while non-normal computerized tomography (CT) scan (OR 15.4, 95% CI 2.2-100+, p=0.002) was associated with SAE.

**Conclusion:** Severe bandemia was not associated with higher odds of adverse events among the general ED pediatric population. Clinicians should base their clinical judgment on the overall context of history, physical examinations, and other laboratory and imaging data.

## INTRODUCTION

More than 42,000 cases of severe sepsis were reported among children in the United States in 1995, with a mortality rate of about 10%; millions of children throughout the world contract this potentially lethal condition each year^1^. In 1992, the Society of Critical Care Medicine designated a band cell concentration of more than 10% of the total peripheral white blood cell (WBC) count as one of the criteria for systemic inflammatory response syndrome^2^. Since then, many clinicians have considered an elevated band cell level as a surrogate for serious bacterial illness (SBI).

Bandemia was associated with blood stream infection in the adult patient population^3^. In the pediatric emergency medicine literature, a sustained controversy questions whether a band cell level greater than 10% of the WBC count truly indicates SBI. Jaskiewicz et al., writing in 1994, and Schnadower et al., publishing in 2010, claimed that pediatric patients with bandemia greater than 10% were at risk of SBI^4,5^. In contrast, other author groups in the past 15 years reported no association between bandemia greater than 10% and SBI^6–8^. Kuppermann et al. suggested that bandemia at 13% did not differentiate between pediatric patients with culture-proven or laboratory-proven bacterial infections and those with viral infections^8^.

Most published studies considered a band level of 10% as the threshold for bacterial infection. There is scant evidence in the pediatric emergency medicine literature about the association between severe bandemia (>20% band cells) and clinical outcomes. We hypothesized that severe bandemia is not associated with an increased risk of adverse events and outcomes.

## METHODS

### Study Design

We conducted a retrospective cohort study using medical records from a single pediatric emergency department (ED) at an urban, regional tertiary referral level 1 pediatric trauma center. The study was approved by the hospital’s institutional review board.

### Patient Population & Data Collection

We created a convenience cohort by first identifying patients who were 2 months to 18 years of age when they were brought to the ED for evaluation between January and July 2008. We selected this date because our regional pediatric hospital ceased to perform differential cell count on all patients’ blood samples. Starting on August 01, 2008, differential cell count, and band levels, were only performed by orders of physicians who suspected infection in their patients. As a result, blood samples with bands after August 01, 2008 were most likely performed on patients with high suspicion for infection.

Severe bandemia was defined as band cell levels ≥20 (constituting 20% of the total peripheral WBC count)^9^. Any imaging study (computerized tomography [CT] scan or X-ray [XR] films) interpreted by attending radiologists as suggesting an infectious process would be considered “non-normal” imaging studies.

An investigator who was not blind to the study hypothesis abstracted the data into a standardized form using an Excel spreadsheet (Microsoft Corporation, Redmond, WA). Data was subsequently prepared by other investigators who were blind to the study hypothesis prior to analysis.

### Outcomes

The primary outcome in this study was a severe adverse event (SAE) associated with bacterial illness. SAEs were defined as in-hospital death, hospital admission from the index ED visit or within 7 days, returning to the ED within 7 days, bacteremia (blood culture showing >10,000 colony-forming units per milliliter), or positive cerebrospinal fluid (CSF WBC count >10 cells per high-power field, positive gram stain).

The secondary outcome was any moderate adverse event (MAE) associated with bacterial illness. These events were defined as positive urinalysis (urinary WBC count >100,000 cells/high-power field, positive nitrite or leukocyte esterase); bacteria-positive stool cultures. Other outcome was any adverse event (AAE), which is a combination of SAEs and MAEs.

### Data Analysis

We first performed descriptive analyses to categorize patients in each subgroup. Continuous variables were assessed for normal distribution by the Ryan-Joiner test. Normally-distributed-data was expressed as mean (standard deviation [SD]) and compared via the Student’s t-test. Chi-square tests were used to compare groups of categorical data, while Analysis of Variance (ANOVA) with Holm-Sidak post-hoc tests, were used to compare means between groups of normally-distributed-continuous-data.

We also performed multivariable logistic regressions to assess association between clinical factors and outcomes (SAE, MAE, or AAE). Statistical analyses were performed with Sigma Plot version 13.0 (Systat Software Inc., San Jose, CA). Two-tailed p-values <0.05 were considered significant.

## RESULTS

The records of 113 patients treated during the 6-month study period indicated severe bandemia. We excluded 11 patients from that original group: 4 because the measured band level was missing from the record, and seven because they were over 19 years of age; therefore, 102 patients were included in the final analysis. Mean (SD) was 5.25 (0.5) years.

Twenty-one patients (21%) had SAEs, 18 (18%) had MAEs, 39 (38%) AAE (**Table 1**), and 63 (62%) patients did not have AAE.

**Table 1.**
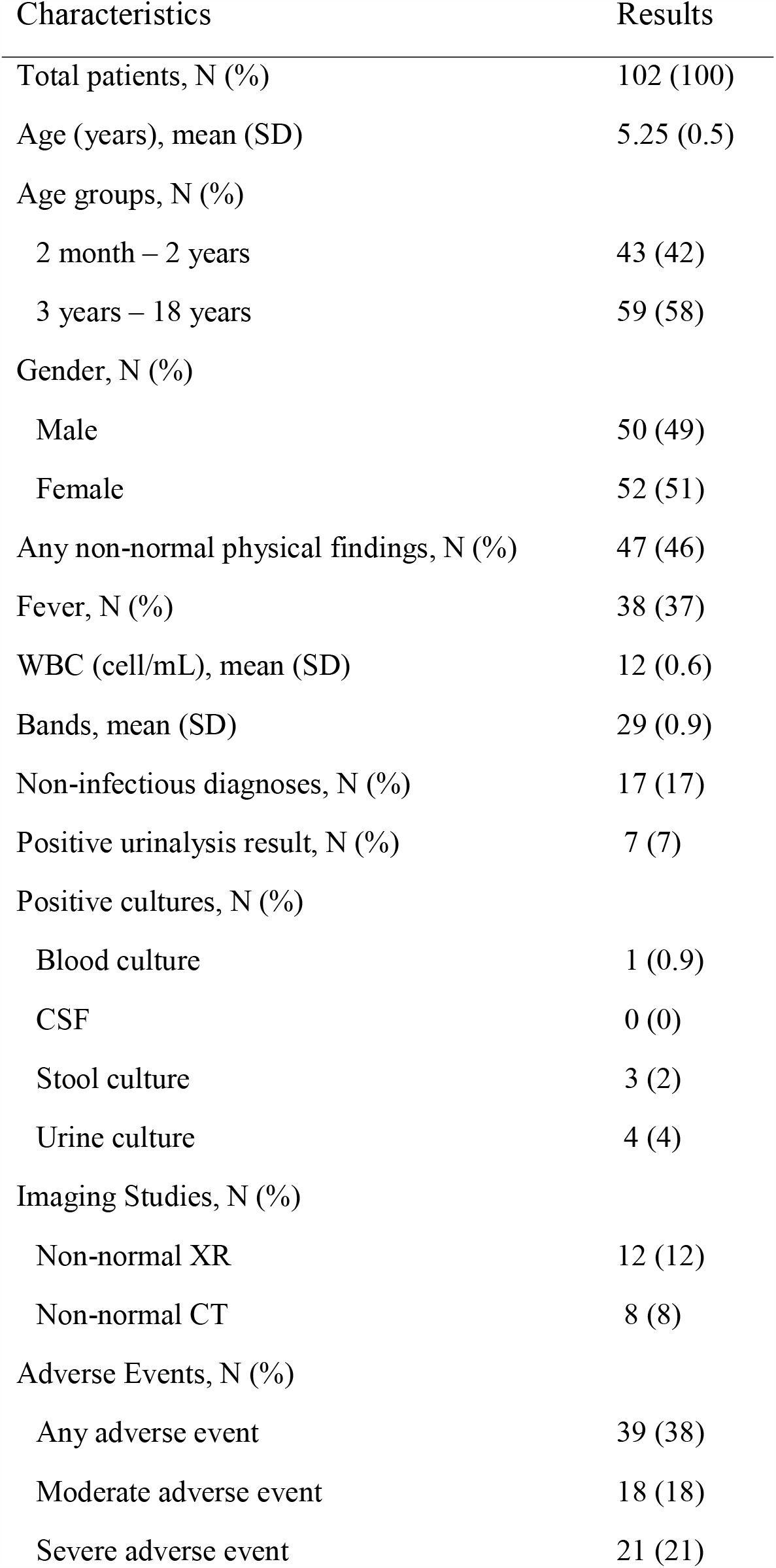
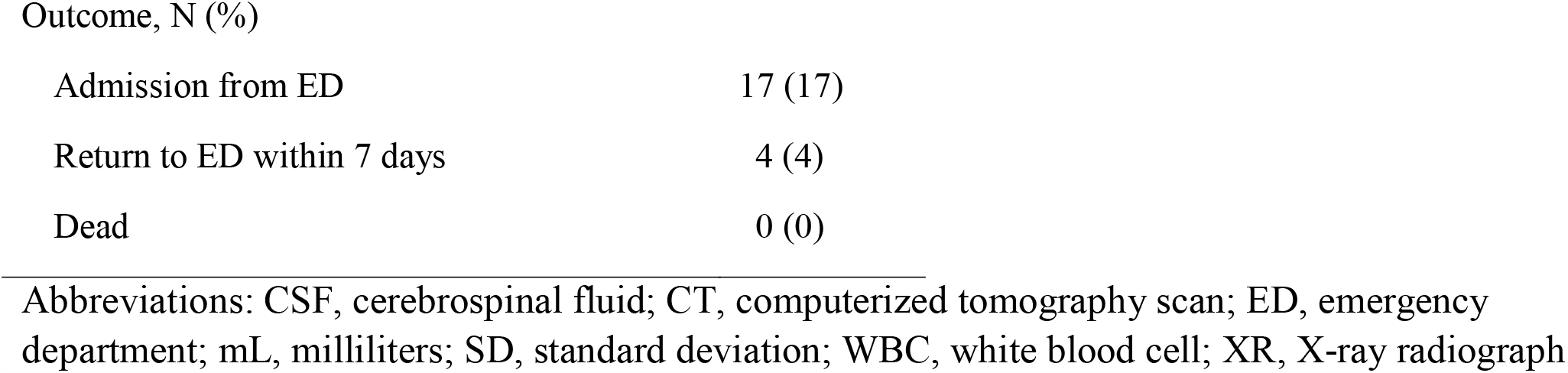
Characteristics of pediatric patients with severe bandemia. Bands were counted as percentage of total white blood cell (WBC) count.

| Characteristics | Results |
| --- | --- |
| Total patients, N (%) | 102 (100) |
| Age (years), mean (SD) | 5.25 (0.5) |
| Age groups, N (%) |  |
| 2 month – 2 years | 43 (42) |
| 3 years – 18 years | 59 (58) |
| Gender, N (%) |  |
| Male | 50 (49) |
| Female | 52 (51) |
| Any non-normal physical findings, N (%) | 47 (46) |
| Fever, N (%) | 38 (37) |
| WBC (cell/mL), mean (SD) | 12 (0.6) |
| Bands, mean (SD) | 29 (0.9) |
| Non-infectious diagnoses, N (%) | 17 (17) |
| Positive urinalysis result, N (%) | 7 (7) |
| Positive cultures, N (%) |  |
| Blood culture | 1 (0.9) |
| CSF | 0 (0) |
| Stool culture | 3 (2) |
| Urine culture | 4 (4) |
| Imaging Studies, N (%) |  |
| Non-normal XR | 12 (12) |
| Non-normal CT | 8 (8) |
| Adverse Events, N (%) |  |
| Any adverse event | 39 (38) |
| Moderate adverse event | 18 (18) |
| Severe adverse event | 21 (21) |
| Outcome, N (%) |  |
| Admission from ED | 17 (17) |
| Return to ED within 7 days | 4 (4) |
| Dead | 0 (0) |

Abbreviations: CSF, cerebrospinal fluid; CT, computerized tomography scan; ED, emergency department; mL, milliliters; SD, standard deviation; WBC, white blood cell; XR, X-ray radiograph

Among the 21 SAE patients, 17 were admitted during the index ED visit and 4 were discharged during the initial visit but returned to the ED within 7 days (**Table 1**). There was no mortality and there was no readmission within 7 days after the index ED visit. Most common diagnosis for 39 patients with AAE was pneumonia (10%), fever (5%), and appendicitis (4%) (**Figure 1**).

**Figure 1.**
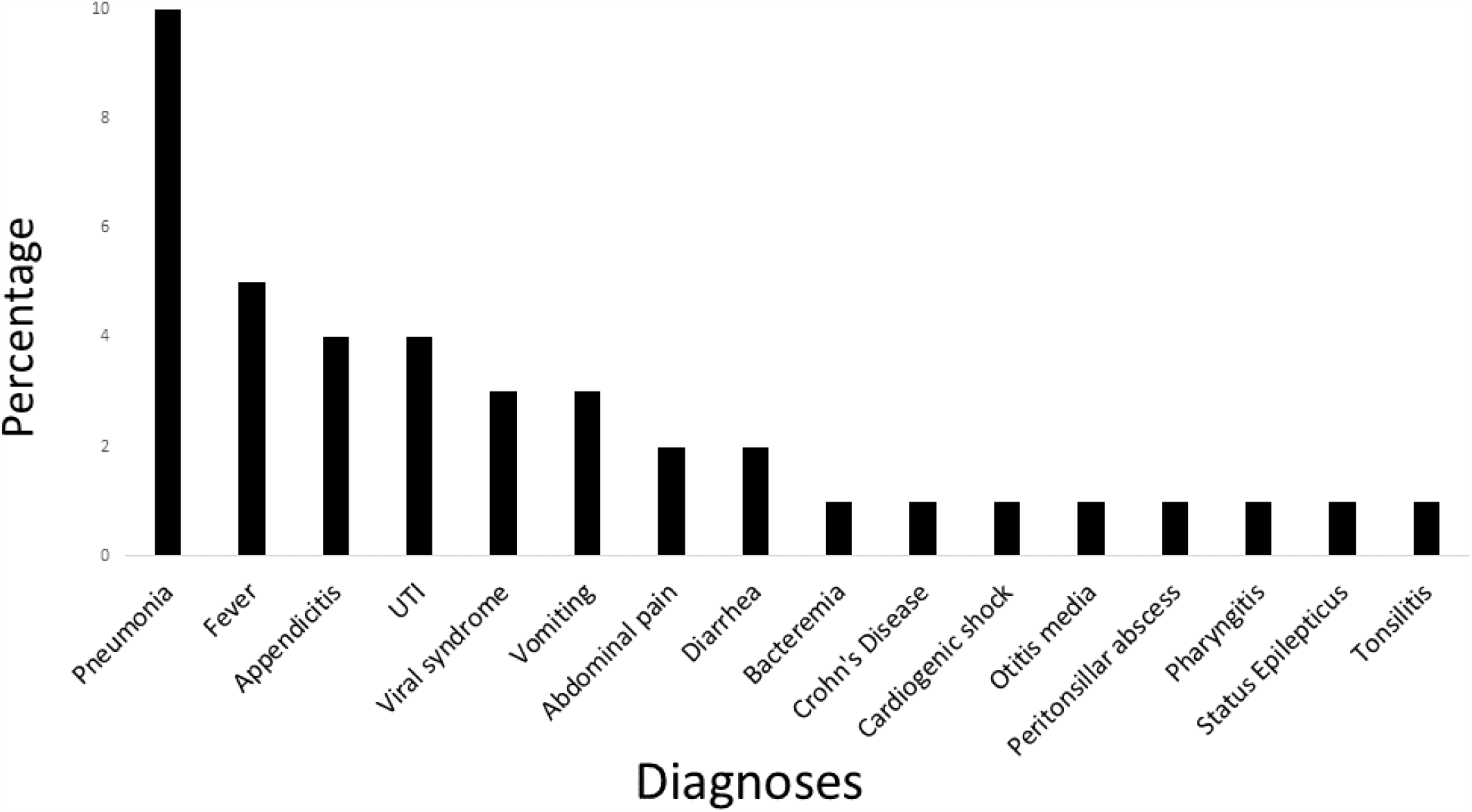
Hospital diagnoses of patients who had any adverse events Abbreviations: UTI, urinary tract infection

Characteristics of patients between groups were similar (**Table 2**). Mean band levels for patients with no adverse events was 28 (10). This level was non-statistically similar (p-value = 0.64) to patients with SAE [31 (9)], MAE [27 (8)], or AAE [29 (9)]. Patients with no adverse events had significantly less non-normal XR and CT findings (p<0.001), compared to those with adverse events (**Table 2**).

**Table 2.** Characteristics of patients having adverse events. *Bands were defined as percentage of total white blood cell (WBC) count

| Variables | No Adverse Events<br>(N=63) | SAE<br>(N=21) | MAE<br>(N=18) | AAE<br>(N=39) | *P* |
| --- | --- | --- | --- | --- | --- |
| Age (years), mean (SD) | 5 (5) | 7 (5) | 4 (4) | 6 (5) | 0.14 |
| Gender, N (%) |  |  |  |  | 0.35 |
| Female | 31 (49) | 14 (66) | 7 (39) | 21 (54) |  |
| Male | 32 (50) | 7 (33) | 11 (61) | 18 (46) |  |
| Non-normal PE findings, N (%) | 29 (46) | 13 (62) | 5 (28) | 18 (4) | 0.21 |
| Fever, N (%) | 24 (48) | 5 (24) | 9 (50) | 14 (36) | 0.40 |
| WBC ($10^3$ cell/mL) | 11 (5) | 13 (8) | 14 (6) | 14 (7) | 0.07 |
| Bands*, mean (SD) | 28 (10) | 31 (9) | 27 (8) | 29 (9) | 0.64 |
| Non-normal XR finding, N (%) | 1 (2) | 3 (14) | 8 (44) | 11 (28) | <0.001 |
| Non-normal CT finding, N (%) | 1 (2) | 6 (28) | 1 (5) | 7 (18) | 0.002 |

Abbreviations: AAE, any adverse event; CT, computerized tomography scan; mL, milliliters; MAE, moderate adverse event; PE, physical examination; SAE, severe adverse event; SD, standard deviation; WBC, white blood cell; XR, X-ray radiograph

Multivariate logistic regressions, after adjusted for age, gender, physical examinations, presence of fever, WBC, bands, XR and CT results, showed that band levels were not associated with SAE (OR 1.04, 95% CI 0.9-1.1, p-value=0.27), MAE (OR 0.97, 95% CI 0.9-1.1, p-value=0.43), or AAE (OR 1.01, 95% CI 0.9-1.1, p-value=0.49) (**Table 3**). Non-normal XR was significantly associated with MAE (OR 17, 95% CI 3.3-90, p-value<0.001) and AAE (OR 32, 95% CI 3.5-100+, p-value<0.001). Non-normal CT was associated with SAE (OR 15, 95% CI 2.2-100+, p-value=0.002) and AAE (OR 27, 95% CI 2.5-100+, p-value=0.001).

**Table 3.** Results from multivariate logistic regressions.

| Variables | AAE |  |  | MAE |  |  | SAE |  |  |
| --- | --- | --- | --- | --- | --- | --- | --- | --- | --- |
|  | OR | 95% CI | *P* | OR | 95% CI | *P* | OR | 95% CI | *P* |
| Age (years) | 0.97 | 0.8-1.1 | 0.62 | 0.97 | 0.8-1.1 | 0.58 | 1.03 | 0.9-1.2 | 0.61 |
| Gender | 0.91 | 0.3-2.6 | 0.85 | 0.91 | 0.2-2.5 | 0.42 | 0.47 | 0.1-1.5 | 0.20 |
| Non-normal PE | 0.84 | 0.3-2.4 | 0.76 | 0.84 | 0.3-2.4 | 0.16 | 1.15 | 0.4-3.7 | 0.82 |
| Fever | 0.68 | 0.2-2.2 | 0.52 | 0.68 | 0.2-2.2 | 0.92 | 0.84 | 0.2-3.1 | 0.80 |
| WBC | 1.04 | 0.9-1.2 | 0.38 | 1.01 | 0.9-1.1 | 0.82 | 1.03 | 0.9-1.1 | 0.45 |
| Bands | 1.01 | 0.9-1.1 | 0.49 | 0.97 | 0.9-1.1 | 0.43 | 1.04 | 0.9-1.1 | 0.27 |
| Non-normal XR | 32 | 3.5-100+ | <0.001 | 17 | 3.3-90 | <0.001 | 1.08 | 0.2-1.2 | 0.94 |
| Non-normal CT | 27 | 2.5-100+ | 0.001 | 1.2 | 0.1-14 | 0.91 | 15.4 | 2.2-100+ | 0.002 |

Abbreviations: AAE, any adverse event; CI, confidence interval; CT, computerized tomography scan; MAE, moderate adverse event; OR, odds ratio; PE, physical examination; SAE, severe adverse event; WBC, white blood cell; XR, X-ray radiograph

## DISCUSSION

In our study, based on a general pediatric ED population, severe bandemia (band cells constituting more than 20% of the total peripheral WBC count) was not associated with a higher risk of adverse outcomes caused by bacterial illness. None of the patients in our study group died during hospitalization or required hospital readmission within 7 days after their initial ED evaluation. Our observations suggested that severe bandemia should not be used as the single factor that influences clinical decisions.

Further studies in pediatric patients are needed to confirm our finding that severe bandemia was not associated with clinical outcomes. However, our finding was consistent with previous studies involving adult ED patients. Ward et al.^10^ reported that median bandemia levels for septic adult survivors were 9%, compared to 17% for septic non-survivors (p-value=0.766) and increasing band level was not associated with increased odds of death (95% CI 0.99-1.028, p-value=0.354)^10^.

Severe bandemia was noted in patients with other inflammatory causes for admission, such as status epilepticus and cardiogenic shock from congenital heart disease, suggesting that bandemia serves as a good inflammatory marker but might not be a good marker for severe infection or adverse clinical outcomes. In contrast, in a study of children with appendicitis^11^, Whyte and associates reported that nonoperative management failed more often in those with bandemia greater than 11% than in those with bandemia less than 10%, which resulted in longer hospital LOS (17 vs. 9 days). The difference between our findings and Whyte et al.’s could be explained by the low prevalence of appendicitis (4%) in our study population or a low prevalence of SBI among our ED pediatric population with severe bandemia.

Four (4%) patients in the SAE group returned to the ED within 7 days after their initial ED visit because of abdominal pain, vomiting, or fever. None were re-admitted to the hospital. This rate of ED return is similar to those documented in previous reports, i.e., 3% at 48 hours^12,13^ and 6% at 14 days after the index ED visit^13^. Most pediatric patients return to EDs because their disease progresses, and up to 30% of them are admitted^14^. In our population, returning patients with SAE were not admitted at the second visits, possibly because the sample of returning patients was small (4%) and because of the poor association between severe bandemia and these patients’ diseases.

Our study also found no association between severe bandemia and a higher risk of MAE caused by bacterial illness. The most common diagnoses for patients in the MAE group were pneumonia (8%) and urinary tract infection (UTI) (4%). Patients in this group were discharged with antibiotics and did not return to the ED or get re-admitted to the hospital within the subsequent 7 days. Thus, without other indicators such as hypoxia, severe bandemia did not affect outcomes among pediatric patients with these infections.

### Limitations

Our study has several limitations. In addition to those inherent to retrospective studies, we did not have an age-matched control group with band counts constituting <20% of the total peripheral WBC count and thus could not compare the rates of adverse events between the study group and a control group. Therefore, we were not able to calculate sensitivity or specificity of severe bandemia as predictors of clinical outcomes. Furthermore, as a single-center study, we might have missed patients who returned to EDs at other hospitals, although this missing rate might be low because the study hospital is the only tertiary referral pediatric hospital in the region.

This study provides information that we hope will be helpful in the design of future investigations of bandemia and outcomes among pediatric patients. Future studies with a higher percentage of surgical patients will provide more conclusive information regarding severe bandemia in medical vs. surgical patients, such as those with appendicitis, which might be associated with higher levels of bandemia and different outcomes from those of medical pediatric patients.

## CONCLUSION

Severe bandemia is an inflammatory marker and is not associated with higher risks of adverse events caused by bacterial illness among ED pediatric patients. Severe bandemia should not be used as a single factor in clinical decisions. Clinicians should base their clinical judgment on the overall context of physical examination findings, other laboratory data, and imaging studies.

## Data Availability

The data is available upon reasonable request.

## Acknowledgement

We thank Dr. Rick Place and Mr. Travis Tracy for their contributions to the manuscript. We also thank Linda J. Kesselring, MS, ELS, for copyediting the manuscript.

## Notes

### Competing Interest Statement

The authors have declared no competing interest.

### Funding Statement

The authors received no funding for data collection or preparation of this manuscript.

### Author Declarations

The study was approved by the ethics committee at the Inova Fairfax Hospital, Falls Church, VA. The decision was to waive any formal consent.

## REFERENCES

1. Watson RS, Carcillo JA. Scope and epidemiology of pediatric sepsis. Pediatr Crit Care Med. 6(3 Suppl):s3–s5.

2. Bone RC, Balk RA, Cerra FB, Dellinger RP, Fein AM, Knaus WA, Schein RM SWACCC. American College of Chest Physicians/Society of Critical Care Medicine Consensus Conference: definitions for sepsis and organ failure and guidelines for the use of innovative therapies in sepsis. Crit Care Med. 2009;136(5 Suppl):e28.

3. Hsueh L, Molino J, Mermel L. Elevated bands as a predictor of bloodstream infection and in-hospital mortality. Am J Emerg Med. Published online 2020:4-7.

4. Jaskiewicz JA, Mccarthy CA, Richardson AC, et al. Febrile Infants at Low Risk for Serious Bacterial Infection -- An Appraisal of the Rochester Criteria and Implications for Management The online version of this article, along with updated information and services, is located on the World Wide Web at□: Pediatrics. 1994;94(3):390–396.

5. Schnadower D, Kuppermann N, Macias CG, et al. Febrile Infants With Urinary Tract Infections at Very Low Risk for Adverse Events and Bacteremia. Pediatrics. 2010;126(6):1074–1083.

6. Isaacman DJ, Shults J, Gross TK, Davis PH, Harper M. Predictors of Bacteremia in Febrile Children 3 to 36 Months of Age. 2000;106(5):977–982.

7. Kanegaye JT, Nigrovic LE, Malley R, et al. Diagnostic Value of Immature Neutrophils (Bands) in the Cerebrospinal Fluid of Children With Cerobrospinal Fluid Pleocytosis. Pediatrics. 2009;123(6):e967–e971.

8. Kuppermann N, Walton EA, Deangelis CD. Immature Neutrophils in the Blood Smears of Young Febrile Children. 1999;153:261–266.

9. Shi E, Vilke GM, Coyne CJ, Oyama LC, Castillo EM. Clinical outcomes of ED patients with bandemia. Am J Emerg Med. 2015;33(7):876–881.

10. Ward MJ, Fertel BS, Bonomo JB, et al. The degree of bandemia in septic ED patients does not predict inpatient mortality. Am J Emerg Med. 2012;30(1):181–183.

11. Whyte C, Levin T, Harris BH, Whyte C, Levin T HB. Early decisions in perforated appendicitis in childrenLJ: lessons from a study of nonoperative management. J Pediatr Surg. 2008;43(8):1459–1463.

12. Alessandrini EA Grenfell SM, Jacobstein CR, Shaw KN. LJM. Return visits to a pediatric emergency department. Pediatr Emerg Care. 20(3):166–171.

13. Zimmerman DR, McCarten-Gibbs KA, DeNoble DH, et al. Repeat pediatric visits to a general emergency department. Ann Emerg Med. 1996;28(5):467–473.

14. Ali AB Howell J, Malubay SM. PR. Early pediatric emergency department return visits: a prospective patient-centric assessment. Clin Pediatr (Phila). 2012;51(7):651.

